# Heart rate fragmentation improves general anesthesia state classification using machine learning

**DOI:** 10.1101/2025.01.07.25320157

**Authors:** Clément Fauchereau, Fanny Carimalo, Axelle Merienne, Marc Laffon, Emmanuel Godat, Jean-Christophe Aude

## Abstract

Accurate assessment of consciousness during general anesthesia is crucial for optimizing anesthetic dosage and patient safety. Current electroencephalogram-based monitoring devices can be inaccurate or unreliable in specific surgical contexts (*e*.*g*. cephalic procedures). This study investigated the feasibility of using electrocardiogram (ECG) features and machine learning to differentiate between awake and anesthetized states. A cohort of 48 patients undergoing surgery under general anesthesia at the Tours hospital was recruited. ECG-derived features were extracted, including spectral power, heart rate variability and complexity metrics, as well as heart rate fragmentation indices (HRF). These features were augmented by a range of physiological variables. The aim was to evaluate a number of machine learning algorithms in order to identify the most appropriate method for classifying the awake and anesthetized states. The gradient boosting algorithm achieved the highest accuracy (0.84). Notably, HRF metrics exhibited the strongest predictive power across all models tested. The generalizability of this ECG-based approach was further assessed using public datasets (VitalDB, Fantasia, and MIT-BIH Polysomnographic), achieving accuracies above 0.80. This study provides evidence that ECG-based classification methods can effectively distinguish awake from anesthetized states, with HRF indices playing a pivotal role in this classification.

**Author summary:** General anesthesia monitoring is critical for optimizing patient safety and outcomes. While electroencephalogram (EEG)-based systems are commonly used, they have limitations in accuracy and applicability, particularly in cases where EEG electrodes placement is challenging or impossible, such as during cephalic surgeries or when patients have forehead skin lesions. Here, a novel approach using electrocardiogram (ECG) signals and machine learning techniques was used to differentiate between awake and anesthetized states during surgery. A total of 48 patients undergoing surgical procedures under general anaesthesia at the Tours hospital were selected for inclusion in the study. This investigation focused on heart rate fragmentation indices, metrics designed for assessing biological versus chronological age, derived from ECG recordings. The gradient boosting algorithm demonstrates performance comparable to leading methods reported in the literature for this classification task. Importantly, model generalizability was confirm through successful application to publicly available datasets. This article highlights the potential of ECG signals as an alternative source for deriving depth of anesthesia indices, offering increased versatility in clinical settings where EEG monitoring is challenging or contraindicated.

## Introduction

General anaesthesia (GA) aims to induce a temporary loss of consciousness in patients undergoing a surgical procedure. This technique is considered a safe procedure, however potential risks and complications may arise [1, 2]. Anesthetic agents affect the function of the brain and the central nervous system, which can result in a range of side effects and complications [3]. During surgery, it is essential to monitor the depth of anesthesia to differentiate between excessive, adequate, or possibly insufficient narcosis states. This will guide the administration and adjustment of volatile or intravenous anaesthetic agents. Correctly dosed hypnotic agents limit haemodynamic morbidity and improve the quality and speed of recovery [4, 5]. Misjudging the depth of anesthesia can be dangerous, especially in children, frail or elderly patients. Underdosing anesthesia can lead to intraoperative consciousness and psychological trauma, such as post-traumatic stress disorder. Conversely, an overdose of anaesthetic can result in a deeper state of anesthesia, which can cause tissue shock, leading to organ failure, brain damage, cognitive dysfunction or postoperative delirium. In extreme cases, very high concentrations of anesthetics drugs can be life-threatening [6]. Accurately measuring the depth of anesthesia is crucial in this context, especially for at-risk patients, to minimize surgical complications. In addition, surgical procedures necessitating GA are expected to increase due to the rising incidence of chronic disorders such as diabetes, cardiovascular disease, cancer, stroke and kidney disease.

Brain activity monitoring devices are necessary to support practitioners in assessing the depth of anesthesia, as clinical assessment alone (*e*.*g*. Guedel classification) is often insufficient. Electroencephalogram (EEG)-based monitors have become the *de facto* standard. The raw EEG signal is transformed into anaesthetic depth indices using proprietary algorithms [7], such as the bispectral index (BIS) (Medtronic, Minneapolis MN, USA), E-entropy (GE Healthcare, Chicago IL, USA), Narcotrend Index (Monitor Technik, Bad Bramstedt, Germany), or patient state index (PSI) (Masimo, Irvine CA, USA). The BIS is the most widely used and the first to be approved by the United States FDA. Several studies have demonstrated its ability to improve anaesthetic dosing, reduce cases of short anesthesia and improve recovery [8]. Various methodological approaches have been described in the literature using EEG to predict the depth of anesthesia, such as frequency and/or time domain analysis [9], artificial neural networks [10], entropy or complexity measures [11], multi-scale methods [12], Poincaré analysis [13], convolution networks or deep learning [14].

Yet, accurately estimating the depth of anesthesia remains a challenge due to various factors such as the use of numerous different drugs (*e*.*g*. opioids, paralytics…), the presence of comorbidity (*e*.*g*. cardiovascular diseases, diabetes…), and inter-patient variability. Additionally, EEGs signals are susceptible to artifacts arising from electromagnetic interference within the operating room environment (*e*.*g*. electric scalpels, powerlines) and electrode displacement, leading to a diminished signal-to-noise ratio. It should also be noted that EEG monitoring may not be appropriate for some clinical settings, such as during cephalic surgery or when there are skin lesions (*e*.*g*. burns) on the forehead.

During surgeries a systematic monitoring of patients cardiac activity through an electrocardiogram (ECG) is mandatory in most countries for legal reasons. Kanaya *et al*. [15] have shown that some hypnotic agents induce changes in the cardiac system. For instance, heart rate variability (HRV), which is associated with the regulation of the autonomic system, is significantly impacted by GA and varies depending on the anaesthetic agents administered [16] and have long been described in the literature as a means of detecting different phases of consciousness [17]. Additionally, it has been demonstrated that some anaesthetic agents, particularly propofol, also affect the time taken for ventricular depolarisation and repolarization (*i*.*e*. QT interval) [18].

Numerous studies have demonstrated the potential of ECG signals for estimating the depth of anesthesia. These investigations typically employ signal processing techniques to filter and to remove noise on raw ECG data before identifying the PQRST wave complex using specialized algorithms. For instance, Martinez J. *et al*. [19] developed a robust algorithm, based on wavelets decomposition, for this purpose. Subsequently, researchers extract various features from the delineated ECG signal, with HRV indices derived from the R-R signal (time interval between two R waves) being prominent features of interest. Differentiating anesthesia states can be achieved using various methods operating at different scales. These approaches encompass ECG signal frequency analysis, as well as machine learning and statistical algorithms [20, 21]. Polk S.L. *et al*. [22] used a set of features derived from frequency spectrum analysis, including total power, low frequencies (0.04–0.17 Hz), high frequencies (0.17–0.32 Hz), and temporal features (R-R interval, HRV). These authors used three machine learning algorithms: K-nearest-neighbor, logistic regression, and decision trees to predict states of consciousness (awake or under the effect of anesthetic agents, in particular sevoflurane). Similarly, Zhan J. *et al*. [23] adopted a comparable feature selection strategy, incorporating high-frequency (HF) power, low-frequency (LF) power, the HF/LF power ratio, and signal complexity measured using the sample entropy method [5]. They subsequently implemented a neural network to distinguish among three distinct anesthesia states: induction (transition from wakefulness to anesthesia), maintenance (stable anesthetic state), and recovery (transition from maintenance back to wakefulness).

This article presents a novel method for predicting patient consciousness states during GA using machine learning algorithms and cardiac signals alongside physiological data (*e*.*g*. age, weight). A cohort of 48 patients admitted for surgery was assembled for this study, as detailed in the Methods section. Our findings demonstrate that incorporating heart rate fragmentation (HRF) indices [24] enhances the accuracy of models utilizing ECG frequency and HRV measures. Furthermore, we assessed the generalizability of these models by evaluating them against established reference databases: Fantasia for the awake state [25], MIT-BIH Polysomnographic for the asleep state [26], and VitalDB for the anesthetized state [27].

## Methods

### Study cohort

This study received ethical approval from the Tours hospital bioethics committee (record number 2020–110) prior to enrollment and was validated by the French national data protection comission (“Commission Nationale de l’Informatique et des Libertés”, CNIL). Participants enrolled in this cohort were scheduled for elective surgical procedures at Tours university hospital between November 2021 and November 2022. Inclusion criteria were met by patients aged 18 or older who provided informed consent for participation. Exclusion criteria included patients aged 17 or younger, those who did not provide consent, those under legal protection such as guardianship, those with permanent atrial fibrillation or pacemakers, and those undergoing emergency or cephalic surgeries. Management of GA was left to the discretion of the anesthesiologist in charge of the room, an carried out according to current recommendations of the “Société française d’anesthésie-réanimation” (SFAR). Standard induction included pre-oxygenation with 100 % oxygen, intravenous administration of sufentanil (0.2 µg/kg), ketamine (0.2 to 0.4 mg/kg). Lidocaine (1 to 1.5mg/kg) was sometimes administered. Propofol (2–4 mg/kg) was injected when the expired fraction of oxygen went over 90 %. Either atracurium (0.5 mg/kg), rocuronium (0.6 to 1.2 mg/kg) or local lidocaine pulverizations on glottis were administered either at the clinical loss of consciousness or during the propofol injection. During surgery, unconsciousness is maintained within a BIS target range of 40-60 using sevoflurane as the volatile anesthetic agent.

Electrophysiological signals were recorded in both the pre-operative (awake) and intra-operative (anesthetized) states. Data analysis included only participants with recordings of at least 10 minutes prior to anesthesia induction and a minimum of 35 minutes during surgery, commencing five minutes after induction. Participants with recordings shorter than these criteria were excluded from the analysis. Table 1 provides a comprehensive overview of the characteristics of our cohort of 48 patients.

**Table 1.** Cohort characteristics.

|  |  |  |
| --- | --- | --- |
| **Participants** | 48 |  |
| **Male / Female** | 18 / 30 |  |
| **Age** | 50.60 | ±16.19 |
| **Height (cm)** | 169.04 | ±8.85 |
| **Weight (kg)** | 76.79 | ±13.05 |

### Data acquisition and preprocessing

Physiological signals were acquired using a portable CARESCAPE™ ONE patient monitor connected to a fixed CARESCAPE™ B650 anesthesia monitor. Two data acquisition phases were conducted for each patient, utilizing the same monitor. The initial phase was performed in the preoperative room, with the patient positioned in a supine resting state. The subsequent phase occurred within the operating room, concurrent with the surgical procedure. VitalRecorder software version 1.10.3.0 [27] running on a laptop captured real-time data from both phases. ECG signals were recorded at a sampling rate of 300 Hz and pre-processed to remove artifacts caused by electromagnetic interference using a 5th-order Butterworth bandpass filter, with cutoff frequencies of 0.5 Hz and 100 Hz.

### ECG analysis and segmentation

Fixed length epochs were extracted from each awake and anaesthetized records. While the full length of the awake records were considered, only signal from 5 to 35 minutes after the reported induction time were allowed for anesthetized records. Epochs where randomly sampled in the selected time frames. Records obtained in a surgical setting can contains a lot of artifacts. To minimize the effect of these artifacts, epochs that were considered too noisy to be processed were rejected. The rejection criteria are as follow. First, epochs with missing signal were rejected. Then, the epoch amplitude, calculated as the difference between the maximum and minimum value of the signal was compared to a lower and upper thresholds. The lower threshold of 1 × 10^−5^ mV ensures that the signal is not flat. The upper threshold of 10 mV ensures that the signal does not contains electrode saturation artifacts. To guarantee that the dataset is balanced both in terms of patient and anesthesia state, this successive sampling and rejection process was repeated until the dataset contained 100 epochs per records. The overall *a posteriori* rejection rate was 16.33 % on our dataset. Each epoch was then processed using our implementation of Martinez’ delineation algorithm [19] to detect individual beats in the ECG. This algorithm extracts the peak, onset and offsets of the P, Q, R, S and T waves. It relies on the zero crossing of the wavelet transform to detect peaks in the signal. By considering multiple scales of the wavelet transform, this algorithm is more robust than those relying on the derivative of the ECG signal. The detected R peaks are used to compute the RR signal as the durations between successive R peaks. The obtained RR signal is subsequently corrected using Lipponen’s algorithm [28]. This algorithm detects and corrects or removes abnormal RR values that could distort RR-derived HRV indices.

### Features extraction

This study used a multimodal feature extraction approach using processed ECG signal and patient physiological characteristics indices (see Table 2). ECG-derived features were categorized into five distinct groups:

**Table 2.** Features categorized summary. The first column indicates grouped features by category. The second column is a description of each feature, where NN is the duration between two successive heartbeats.

| Category | Features |
| --- | --- |
| **Morphological metrics**<br>(Yeap T.H. *et al.* [29]) | Q-T, P-R, S-T, Q-R-S interval & segment<br>(mean and standard deviation) |
| **Time domain HRV**<br>(Shaffer F. *et al.* [30]) | standard deviation of NN intervals (SDNN)<br>root mean square of successive NN differences (RMSSD)<br>number of pairs of successive NNs intervals $\geq 50$ ms (NN50)<br>proportion of NN50 divided by total number of NNs (pNN50) |
| **Frequency domain HRV**<br>(Shaffer F. *et al.* [30]) | very low frequencies (VLF) [0.0033–0.04 Hz]<br>low frequencies (LF) [0.04–0.15 Hz]<br>high frequencies (HF) [0.15–0.4 Hz]<br>sympathovagal balance (LF/HF)<br>total power (VLF+LF+HF) |
| **Information dimension**<br>(Bandt C. *et al.* [31], Richman J. *et al.* [32]) | permutation entropy (PermEn)<br>sample entropy (SampEn) |
| **Heart rate fragmentation**<br>(Costa M.D. *et al.* [24]) | percentage of inflection points (PIP)<br>inverse of the average length of accel./decel. segments (IALS)<br>percentage of short segments (PSS)<br>percentage of NN intervals in alternation segments (PAS) |
| **Patient characteristics** | age, height, weight, body mass index (BMI) |

- *Morphological metrics*: quantifying the morphology of ECG signals using statistics on durations between characteristic beat points.
- *Time domain HRV* : quantifying the fluctuations in time intervals between consecutive R-waves.
- *Frequency domain HRV* : analyzing the spectral distribution of ECG signals to identify dominant frequency components.
- *Information dimension*: assessing the inherent non-linearity and unpredictability of ECG signals using entropy metrics.
- *heart rate fragmentation*: probing the degree of sinus rhythm fragmentation of cardiac interbeat interval time series.

### Morphological metrics

Morphological metrics are used to describe the morphology of the segmented beats of an ECG in accordance with the electrical activity of the heart [29]. They measure durations between characteristics points. In the present study, the mean and standard deviation were calculated for each duration series as indicated below.

- *Q-T interval:* from the beginning of the QRS complex to the end of the T wave.
- *P-R interval:* from the beginning of the P wave to the beginning of the QRS complex.
- *P-R segment:* from the end of the P wave to the beginning of the QRS complex.
- *S-T interval:* from the end of the QRS complex to the end of the T wave.
- *S-T segment* from the end of the QRS complex to the beginning of the T wave.
- *Q-R-S interval:* from the beginning to the end of the QRS complex

### Time domain HRV

Time domain HRV is a well established cardiac feature type. These indices aim to quantify the amount of heart rate variability during monitoring [30]. We considered four time domain HRV methods [33]: SDNN, the standard deviation of the interbeat (NN) intervals; RMSSD, the square root of the mean squared differences of successive NNs intervals; NN50, the number of interval differences of successive NN intervals greater than 50 ms; and pNN50, the proportion derived by dividing NN50 by the total number of NN intervals.

### Frequential HRV

We also considered frequential HRV methods [33]. From the power spectral density we derived the power of three frequency bands: VLF, the very low frequency band [0.0033–0.04 Hz]; LF, the low frequency band [0.04–0.15 Hz]; and HF, the high frequency band [0.15–0.4 Hz]. From these frequency band powers, we derived the sympathovagal balance (LF/HF) and the total power (VLF+LF+HF).

### Information dimension

Information dimension methods provide a quantitative framework for assessing the complexity and predictability in time series data. These methods emerged from the field of chaos theory and are particularly suited for analyzing systems exhibiting nonlinear dynamics, such as physiological signals. While encompassing several categories, including capacity dimension, correlation dimension (fractal dimension), Lyapunov dimension, and information dimension (entropy), this study focuses on the latter class due to its demonstrated efficacy in HRV analysis [34]. Two specific entropy metrics were selected for this investigation: PermEn, the permutation entropy [31]; and SampEn, the sample entropy [32], an advancement upon the approximate entropy (ApEn) proposed by Pincus *et al*. [35].

### Heart rate fragmentation

HRF indices aim to estimate the degree of irregularity or *fragmentation* in the ECG signal. These methods analyze the beat-to-beat signal to quantify the number of variations or fluctuations. In this article we use four HRF indices introduced by Costa M.D. *et al*. [24].

Let 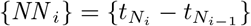 the series of interbeat intervals, where 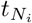 the time of occurrence of the *i*^*th*^ beat, and {Δ*NN*_*i*_} = {*NN*_*i*_ ™ *NN*_*i*™1_} the series of the differences between two consecutive *NN* intervals. Then we derive the series of fluctuations *f*_*i*_ as:

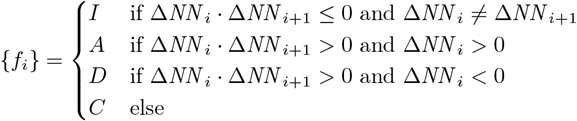

where, *I, A, D, C* are respectively: Inflection (*A*/*D* or *D*/*A* inversions and change to or from *C*), Acceleration, Deceleration and Constant fluctuations. This series depicts accelerations, decelerations or constants successive fluctuation events (segments) separated by inflection points. {*F*_*j*_} is a compacted representation derived from {*f*_*j*_}:

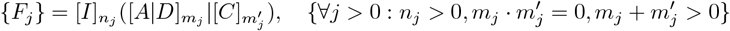

where [*x*]_*n*_ is a succession of *n* ≥ 0 events *x*.

The PIP index, the percentage of inflection points, is defined as:

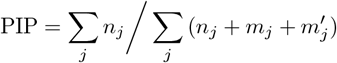

The IALS index, the inverse of the average length of acceleration/deceleration segments, is defined as:

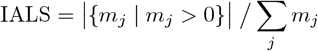

The PSS index, the percentage of short (less than 3 fluctuations) acceleration or deceleration segments, is defined as:

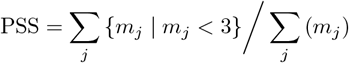

We also consider the series of successive alternation segments defined as:

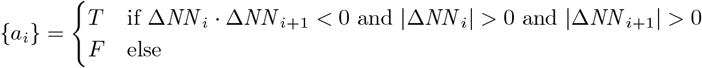

where *a*_*i*_ = *T* (*i*.*e*. True) when two successive *NN* intervals alternate (acceleration to deceleration or conversely) and *a*_*i*_ = *F* (*i*.*e*. False) otherwise. The series {*a*_*i*_} can be transformed in the following compacted representation {*A*_*j*_}:

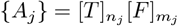

The PAS index, the percentage of NN intervals in alternation segments (sequence of at least four NN intervals), is defined as:

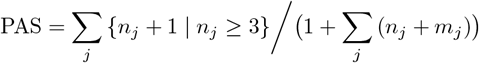

### Models and performance evaluation

In this study we evaluated six machine learning methods for the GA classification task (see Table 3): logistic regression, decision tree, random forest, support vector machine (SVM), and gradient boosting (XGBoost). Models performance were assessed using nested cross-validation [36], a method that integrates model selection and hyperparameter tuning to minimize overfitting the training dataset. Hyperparameters (see Table 3) were optimized through random exploration of the parameter space according to the search distributions in Table 3. All parameters, except hyperparameters, are set to their default values within their respective software implementations. See the Software Implementation section for details.

**Table 3.** Machine learning models and their associated hyperparameters. For each model (first column) hyperparameters are indicated in the second column and the associated probability distribution used for hyperparameter search in the third column.

| Model | Hyperparameters | Search distribution |
| --- | --- | --- |
| **Decision Tree** | Maximum depth | $\mathcal{U}[3, 10]$ |
| **Logistic Regression** | Regularization strength | $\mathcal{U}_{log}(0.1, 100)$ |
| **Nearest Neighbors** | Number of neighbors | $\mathcal{U}[3, 15]$ |
| **Neural Network** | Number of hidden layers | 3 or 10 |
|  | Hidden layer sizes | 10 or 100 |
| **Random Forest** | Number of trees | $\mathcal{U}_{log}[10, 400]$ |
| | Maximum number of features | $\mathcal{U}(0.1, 0.9)$ |
| **SVM** | Regularization strength | $\mathcal{U}_{log}(0.1, 100)$ |
| **XGBoost** | Maximum depth of a tree | $\mathcal{U}[2, 10]$ |
| | Minimum child weight | $\mathcal{U}[1, 10]$ |
| | Learning rate | $\mathcal{U}(0, 1)$ |

𝒰[*a, b*] is the discrete uniform distribution between *a* and *b*. 𝒰(*a, b*) is the continuous uniform distribution between *a* and *b*. 𝒰_*log*_[*a, b*] is the discrete log-uniform distribution between *a* and *b*. 𝒰_*log*_(*a, b*) is the continuous log-uniform distribution between *a* and *b*.

Following training on the complete dataset, further analysis was conducted using the optimal model architecture.

In their article, Polk S.L. *et al*. [22] investigated the use of ECG for classifying awake and anesthetic states during GA. To the best of our knowledge, this study constitutes the most closely related work in the current literature. Consequently, we used the findings reported by these authors for comparison with our own results.

Feature importance was assessed using Shapeley values [37], while generalization capabilities were evaluated across publicly accessible datasets. Based on our informations, VitalDB [27] is the sole publicly accessible comprehensive repository of high-resolution physiological signal recordings collected during surgical procedures under GA. The VitalDB dataset used in this work was extracted on April 13^th^, 2023. However, unlike our dataset, VitalDB lacks recordings of resting physiological signals acquired during the preoperative period. VitalDB contains 6388 records of which 2324 are records of GA using propofol as anesthetic agent. It is very large compared to other databases so we randomly sampled 300 records out of the 2324 records. For each record, we considered at most 30 minutes of recording starting at the reported start of anesthesia and ending either at the end of the record, the reported stop of anesthesia or 30 minutes after the start of anesthesia. Unlike the ECG processing used in this study, no signal quality filtering was applied. Thus performances were assessed on potentially noisy signals.

To evaluate model generalization across awake and sleep states, we respectively leveraged the Fantasia Database Version 1.0.0 [25] and the MIT-BIH Polysomnographic Database Version 1.0.0 (excluding annotated awake recording segments) [26]. We used all 40 cases denoting awake states from the Fantasia database. Similarly, all 18 cases denoting asleep states from the MIT-BIH Polysomnographic database were included in our analysis. All ECG records extracted from the three databases were segmented using 50 % overlapping sliding windows of 180 seconds, without any signal quality rejection rule applied.

### Evaluation metrics

Machine learning models were evaluated using four standard classification performance metrics: accuracy, F1 score, precision, and recall. Formal definitions of these metrics is as follows:

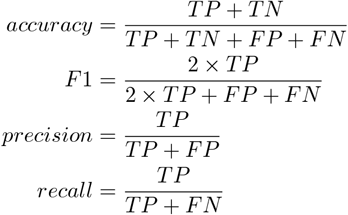

where *TP* is the number of true positives, *TN* is the number of true negatives, *FP* is the number of false positives, and *FN* is the number of false negatives.

Generalization was evaluated by predicting a single label, awake, asleep, or anesthetized, depending on the database used. Due to the absence of true negatives and false positives in this setup, accuracy was deemed the most appropriate metric for evaluating generalization performance.

### Software implementation

Python (version 3.11) and NumPy (version 1.26.4) were used to perform raw data processing, ECG delineation, features calculations, nested cross validation, and data analyses. Machine learning implementations provided by the package Scikit-Learn (version 1.4.0) were used for the following methods: decision tree, logistic regression, nearest neighbors, neural network (multi-layer perceptron), support vector machine (radial kernel), and random forest. The package xgboost (version 2.0.3) was used for gradient boosting algorithm (XGBoost). We use the SHAP (version 0.46.0) package for Shapeley values calculations and plots. Kruskal-Wallis and Dunn post-hoc tests were performed using the R language (version 4.2.2) and the R ‘DescTools’ package (version 0.99.54).

## Results

### ECG segmentation window size and sampling rate

This analysis used two key ECG segmentation parameters: window size and sampling rate (epochs per awake or anaesthetized states records). Feature metric robustness to signal length was a crucial consideration. For instance, sample entropy exhibits instability for short time series [38], requiring higher sampling rates for reliable estimation. Conversely, using larger window sizes can introduce oversampling artifacts. Our analysis determined that a 180 seconds window size and 100 samples per record is an optimal trade-off. Sample entropy, being highly sensitive to time series length, requires a minimum of 100 data points for robust estimation according to the upper bound definition given by Richman J.S. *et al*. [32] (see S1 Fig). This threshold was adopted as a minimum requirement in this study.

To determine suitable window sizes for ECG segmentation, we calculated the number of interbeat intervals within sliding windows ranging from 30 to 300 seconds, considering an average heart rate range of 40 to 80 beats per minute (see Fig 1A). Windows exceeding 180 seconds consistently yielded a minimum of 100 interbeat intervals, satisfying the threshold for reliable sample entropy estimation.

**Fig 1.**
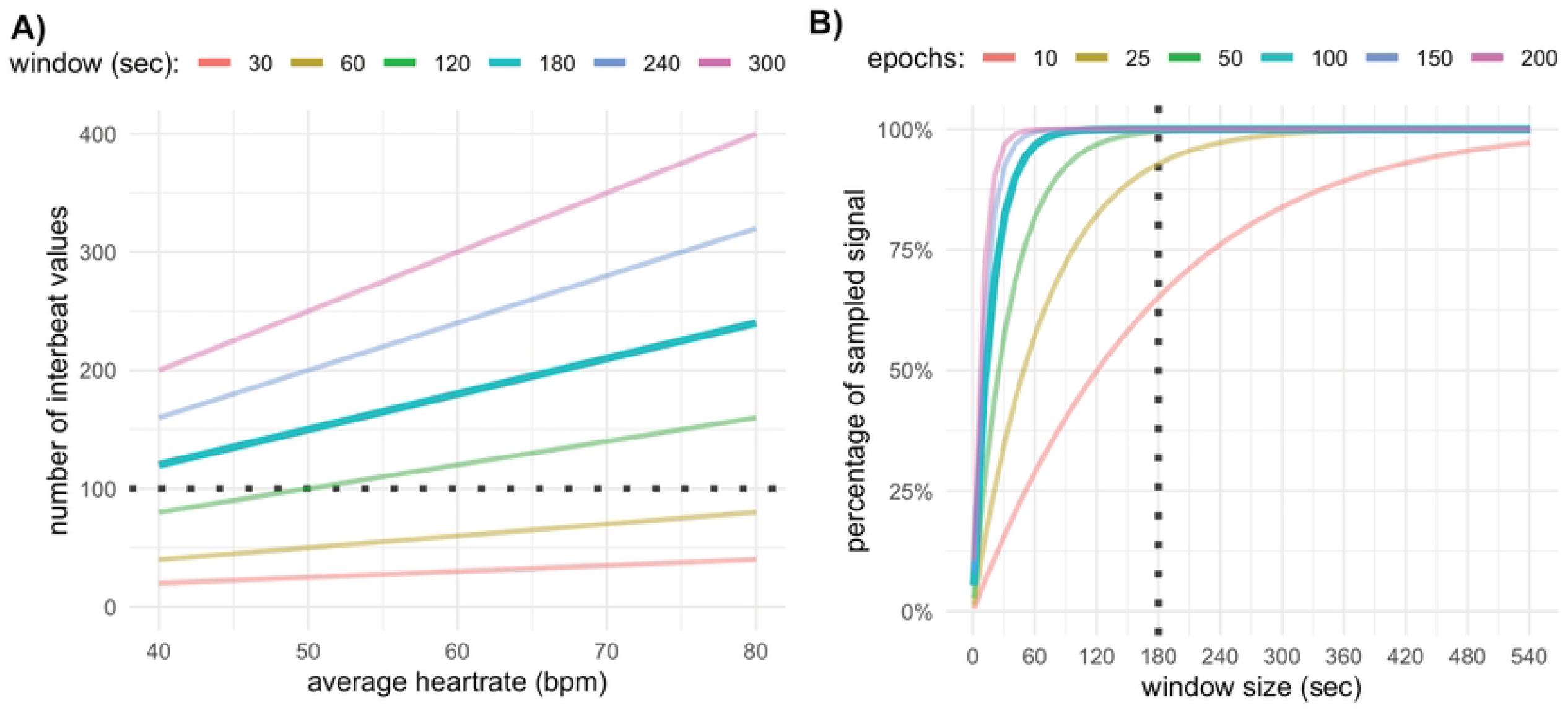
ECG segmentation window sizes and sampling rate. (A) depicts the number of interbeat values (y-axis), for an average heart rate in the range 40 to 80 bpm (x-axis), using colored lines for signal segmentation window of sizes varying from 30 to 300 seconds. The horizontal dotted line depicts the lower bound for the interbeat numbers used in this study. (B) depicts the percentage of sampled ECG signal (y-axis), using a uniform distribution to sample the starting time point of the window. The size of the segmentation window is indicated on the x-axis and each line matches a different sampling rate (number of epochs) varying from 10 to 200. The vertical dotted line depicts the lower bound for the segmentation window size used in this study.

Epochs are sampled using a uniform distribution. The number of sampled epochs required to ensure that every time point of the ECG signal appears in at least one epoch depends on the window size. As illustrated in Fig 1B, window sizes exceeding 180 seconds require at least 100 epochs to ensure complete coverage of the ECG signal under a uniform distribution assumption.

To evaluate the impact of segmentation window size on classification accuracy, we used a support vector machine (SVM) model within a nested cross-validation framework. A sampling rate of 100 epochs was used for this analysis. As depicted in Fig 2, classification accuracy steadily increased with segmentation window size up to 180 seconds, after which further increases yielded minimal improvement. Evaluation with alternative performance metrics led to the same conclusions (see S2 Fig).

**Fig 2.**
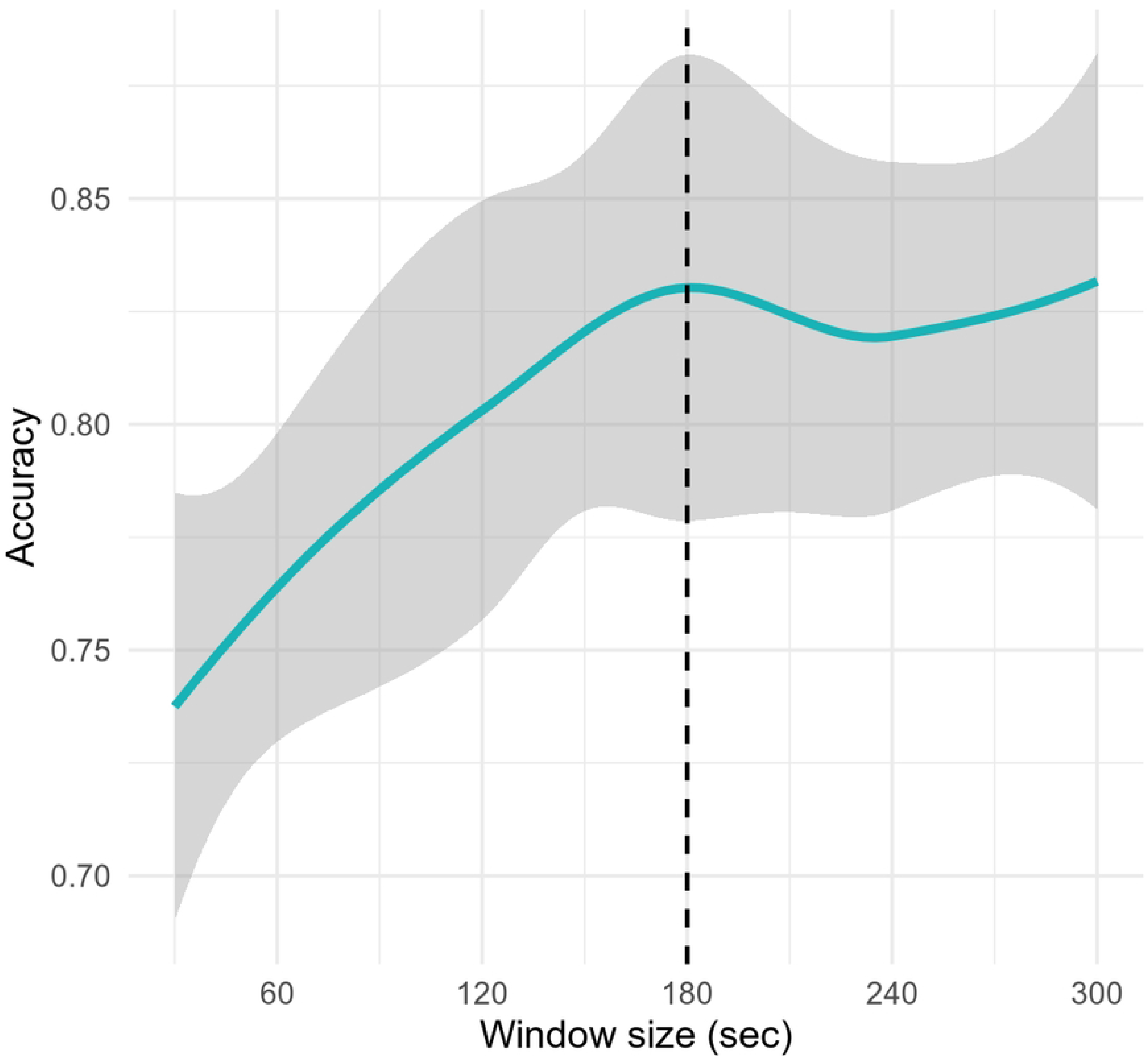
Effect of window size on accuracy for the SVM model. This plot depicts the SVM model accuracy (y-axis), evaluated using nested cross-validation on the whole dataset, for window sizes (x-axis) in the range 30 to 300. For each record of the dataset 100 windows were selected, using a uniform sampling process, amounting to a total of 9600 windows. The solid blue line depicts the average accuracy. The grey region depicts the average accuracy ± the standard deviation. The vertical dashed line shows the chosen window size used in this study.

This analysis confirms that a 3-minutes window size with a sampling rate of 100 epochs represents the optimal configuration for robust ECG segmentation in this study. These parameters will be adopted for all subsequent analyses.

### Model performance

Classification models performance, evaluated using nested cross-validation as described in the Methods section, is presented in Table 4. Decision tree, logistic regression, and nearest neighbors exhibited the lowest performance across all metrics, with scores below 0.8 for all but precision. Notably, decision trees and logistic regression achieved precision scores of 0.83 ± 0.11 and 0.81 ± 0.14 respectively. The neural network model demonstrated slightly improved performance compared to the aforementioned models in all metrics except precision 0.79 ± 0.13.

**Table 4.** Nested cross-validation results. Nested cross-validation results evaluated using models indicated in the first column on the whole dataset. Reported metrics are accuracy, F1 score, precision and recall.

| Model | Accuracy | F1 score | Precision | Recall |
| --- | --- | --- | --- | --- |
| Decision Tree | $0.79 \pm 0.07$ | $0.78 \pm 0.07$ | $0.83 \pm 0.11$ | $0.73 \pm 0.06$ |
| Logistic Regression | $0.79 \pm 0.10$ | $0.78 \pm 0.11$ | $0.81 \pm 0.14$ | $0.76 \pm 0.13$ |
| Nearest Neighbors | $0.77 \pm 0.08$ | $0.76 \pm 0.08$ | $0.76 \pm 0.09$ | $0.78 \pm 0.11$ |
| Neural Network | $0.80 \pm 0.11$ | $0.80 \pm 0.10$ | $0.79 \pm 0.13$ | $0.82 \pm 0.11$ |
| Random Forest | $0.83 \pm 0.08$ | $0.84 \pm 0.06$ | $0.80 \pm 0.11$ | $0.89 \pm 0.05$ |
| SVM | $0.83 \pm 0.10$ | $0.83 \pm 0.10$ | $0.82 \pm 0.13$ | $0.85 \pm 0.12$ |
| XGBoost | $0.84 \pm 0.08$ | $0.84 \pm 0.08$ | $0.83 \pm 0.11$ | $0.86 \pm 0.08$ |

Superior performance was observed with random forest, support vector machine (SVM), and gradient boosting models. XGBoost achieved the highest accuracy 0.84 ± 0.08, F1-score 0.84 ± 0.08, and precision 0.83 ± 0.11. Random forest attained the highest recall score 0.89 ± 0.05.

To contextualize our findings, we compared the achieved F1 score with that reported by Polk S.L. *et al*. [22] (see Table 5). Due to the unavailability of their dataset, a comprehensive comparison was restricted to this metric. It is noteworthy that the anesthetic agents employed in our study differed from those used by Polk S.L. *et al*. Despite these methodological discrepancies, the comparable F1 scores of the best models observed in both studies suggest a similar capacity for differentiating between awake and anesthetized states.

**Table 5.** Performance comparison against existing methods. This table shows a model performances comparison of this study and the one of Polk S.L. *et al*. The dataset is reported in the first column, the analgesic agent(s) used in the second column, the classification model in the third column, and the F1 score in the fourth column.

| Dataset | Anesthetic(s) | Model | F1 Score* |
| --- | --- | --- | --- |
| This study | propofol + sevoflurane | XGBoost | $0.840 \pm 0.008$ |
| | | Logistic Regression | $0.780 \pm 0.011$ |
| | | Nearest Neighbors | $0.760 \pm 0.008$ |
| | | Decision Tree | $0.780 \pm 0.007$ |
| Polk S.L. *et al.* | sevoflurane | Logistic Regression | $0.834 \pm 0.006$ |
| | | Nearest Neighbors | $0.685 \pm 0.009$ |
| | | Decision Tree | $0.784 \pm 0.009$ |
| Polk S.L. *et al.* | sevoflurane + ketamine | Logistic Regression | $0.880 \pm 0.007$ |
| | | Nearest Neighbors | $0.826 \pm 0.010$ |
| | | Decision Tree | $0.682 \pm 0.010$ |

\* in their article, Polk S.L. *et al.* [22], F1 score is the only reported performance metric.

### Generalization performance

To evaluate the robustness, reliability, and generalization capabilities of our models, we assessed the performance of the three best-performing models (random forest, support vector machine, and gradient boosting) on three public databases: Fantasia, VitalDB, and MIT-BIH Polysomnographic (see the Methods section). These datasets depict patients in different physiological states, respectively: awake, anesthetized, and asleep.

The gradient boosting model achieved the best performance across two out of three datasets. Evaluated using the Fantasia dataset (awake state), it outperformed both the random forest and support vector machine models in accuracy scores as reported in Table 6. Evaluated using the VitalDB dataset (anesthetized state), the gradient boosting model also demonstrated superior performance compared to the other two models. However, tested on the MIT-BIH Polysomnographic dataset (asleep state), the random forest model achieved slightly higher accuracy score (0.94) than the gradient boosting model (0.90). Overall the gradient boosting model outperformed the random forest and support vector machine models across different datasets, suggesting its robustness and reliability in classifying physiological states.

**Table 6.** Generalization performances of the random forest, SVM, and XGBoost models. Models were trained on the whole dataset of this study and performances were evaluated using the accuracy metric on the public datasets indicated in the first column.

| Dataset (*consciousness state*) | Random forest | SVM | XGBoost |
| --- | --- | --- | --- |
| Fantasia (*asleep*) | 0.71 | 0.80 | 0.83 |
| VitalDB (*anesthetized*) | 0.77 | 0.73 | 0.80 |
| MIT-BIH Polysomnographic (*awake*) | 0.94 | 0.73 | 0.90 |

The gradient boosting model achieved an accuracy of 0.80 ± 0.08 when tested on our dataset. As shown in Table 6, its performance remained comparable when predicting awake state using the Fantasia database (accuracy=0.83). However, a 4.8 % decrease in accuracy was observed when predicting the anesthetized state using VitalDB. Remarkably, this model exhibited superior performance in predicting the asleep state on the MIT-BIH Polysomnographic database, demonstrating a 7.1 % increase in accuracy compared to its performance on our training dataset.

Fig 3 compares the distributions of the two most discriminative features of the gradient boosting model (see feature importance results below), percentage of inflection points (PIP) and sample entropy, across the three databases. Notably, these plots reveal distinct distribution patterns for each state (awake, asleep, and anesthetized) for both PIP and sample entropy indices. Statistical analysis using the Kruskal-Wallis test yielded highly significant p-values (*p* ≤ 2 × 10 ^−16^) for both feature distributions, indicating a strong difference between GA states. Furthermore, a Dunn post-hoc test confirmed the statistically significant differences (*p*≤ 2 × 10^−16^) between the anesthetized state and both the awake and asleep states.

**Fig 3.**
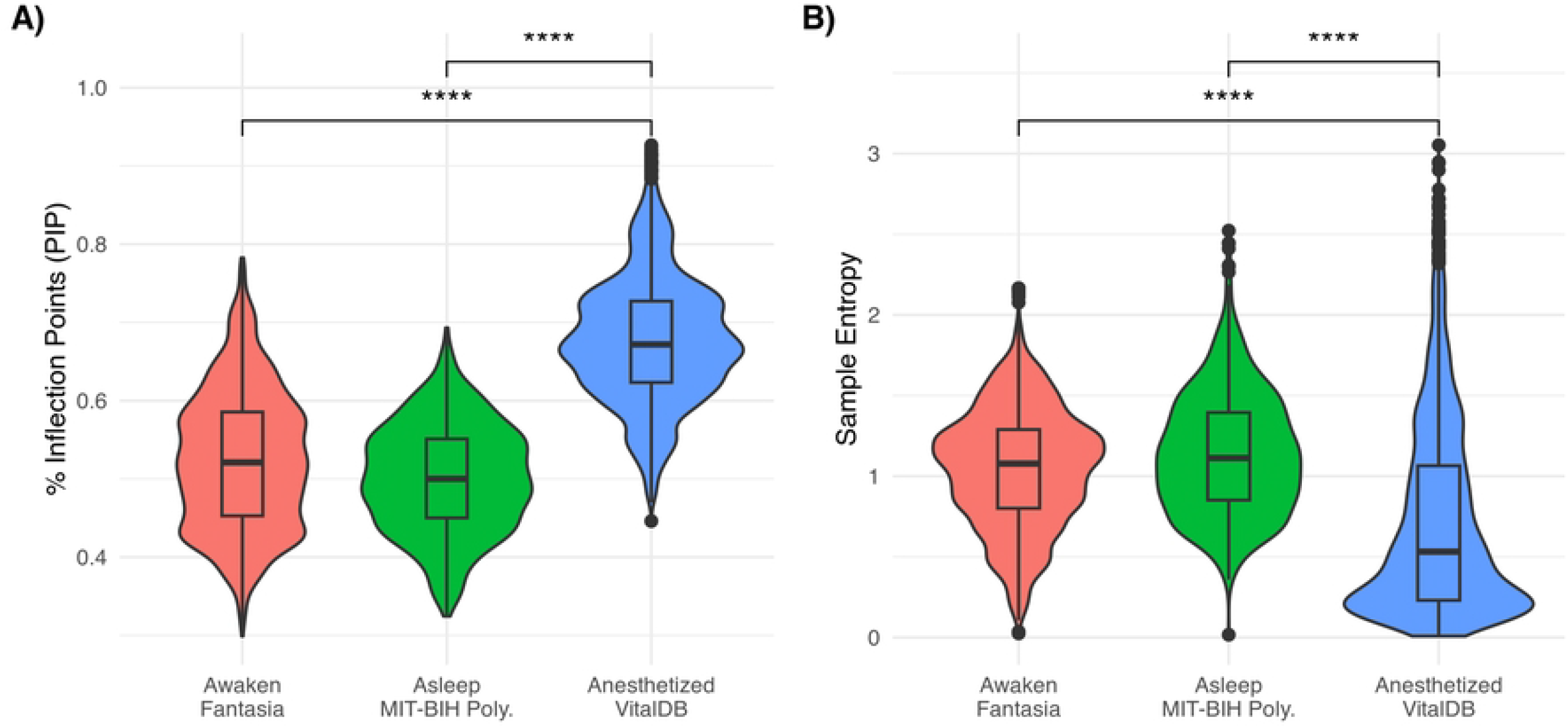
HRF and HRV distributions across consciousness states. (A) This plot depicts distributions of the HRF percentage of inflection points (PIP) index. (B) This plot depicts distributions of the HRV sample entropy (SampEn) index. Distributions are depicted as violin plots for each consciousness state: awake (red) using the Fantasia dataset, asleep (green) using the MIT-BIH Polysomnographic dataset, and anesthetized (blue) using the VitalDB dataset. Windows of 180 seconds were used to calculate HRF and HRV indices. Horizontal segments above distributions indicate statistically highly significant differences between anesthetized states and awake/asleep states as determined by a Dunn post-hoc test (*p* ≤ 2 × 10^−16^).

### Feature Importance

Feature importance was assessed using Shapeley (SHAP) values for the three best-performing models (see Table 4): random forest, support vector machine, and gradient boosting. SHAP values were grouped into six feature categories as detailed in Table 2: morphological metrics, time domain HRV, frequency domain HRV, information dimension, heart rate fragmentation, and patient characteristics. The average and standard deviation of the mean absolute SHAP value were calculated for features within each category. These results, for the gradient boosting model, are shown in Fig 4 (see S3 Fig for SVM and random forest models).

**Fig 4.**
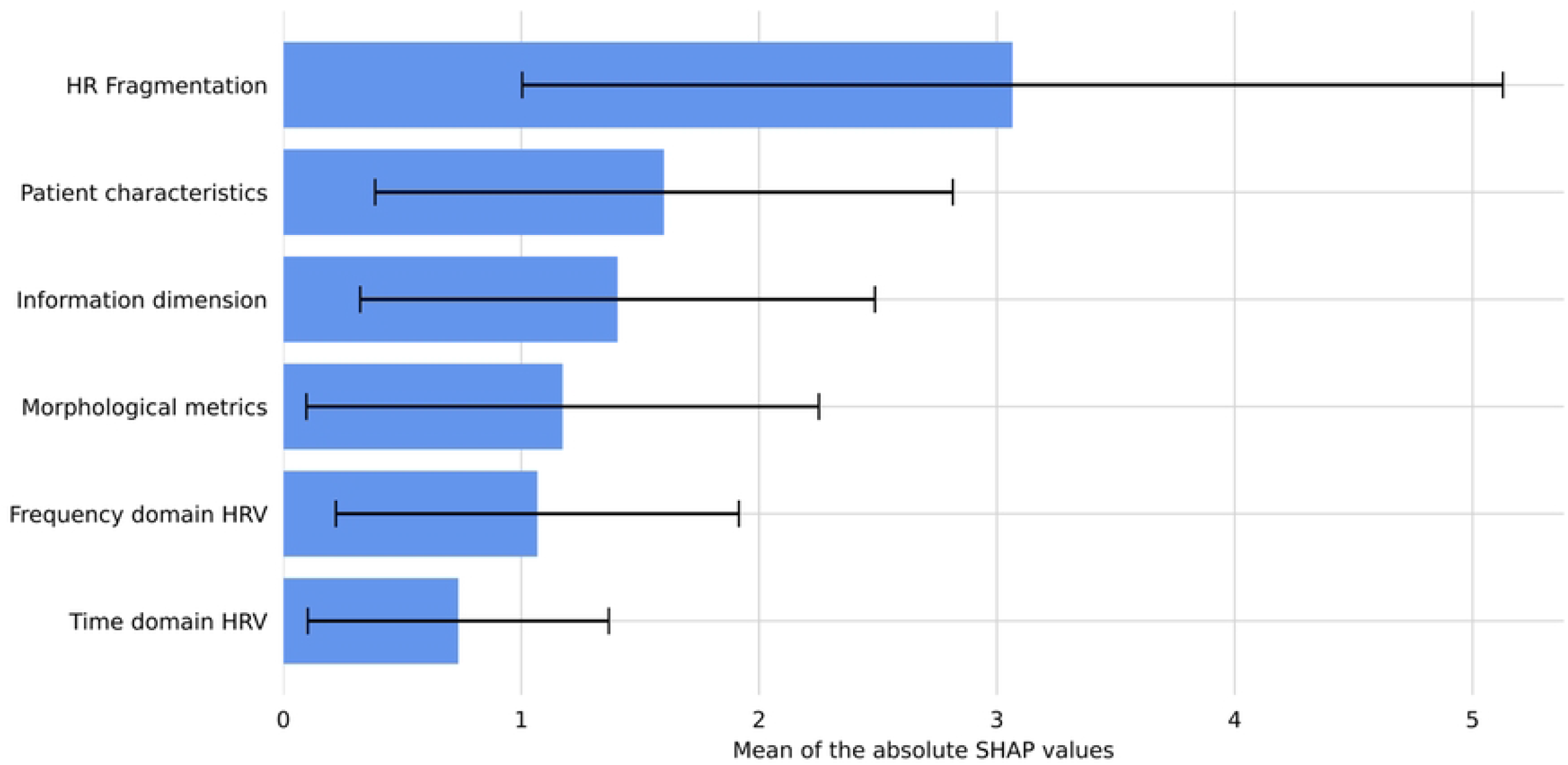
Feature importance grouped by category. This plot depicts the average absolute SHAP values per feature categories. SHAP values were evaluated using the XGBoost model trained on the whole dataset. The standard deviation of the SHAP values is indicated as a black horizontal lines.

Single feature with the highest importance are shown in Fig 5 using a beeswarm plot (see S3 Fig for SVM and random forest models). These plots demonstrate that HRF indices are amongst the most important features to discriminate between awake and anesthetized states. In particular higher values of the percentage of inflection points (PIP), the percentage of alternate segments (PAS), and the inverse of the average length of acceleration or deceleration segments (IALS), respectively first, fourth and ninth on the beeswarm plot Fig 5, have a positive impact on the model predictions (*i*.*e*. to predict the anesthetized state). In order to validate this hypothesis, we also evaluated the three classifiers using two sets of features: without HRF indices, and with only HRF indices and patient characteristics. In the initial model, a decline in performance was observed (see S1 File, Table 1). Conversely, the second model demonstrated a performance level that was comparable to that of the first (see S1 File, Table 2). These results support the contribution of HRF indices to the performances of the model.

**Fig 5.**
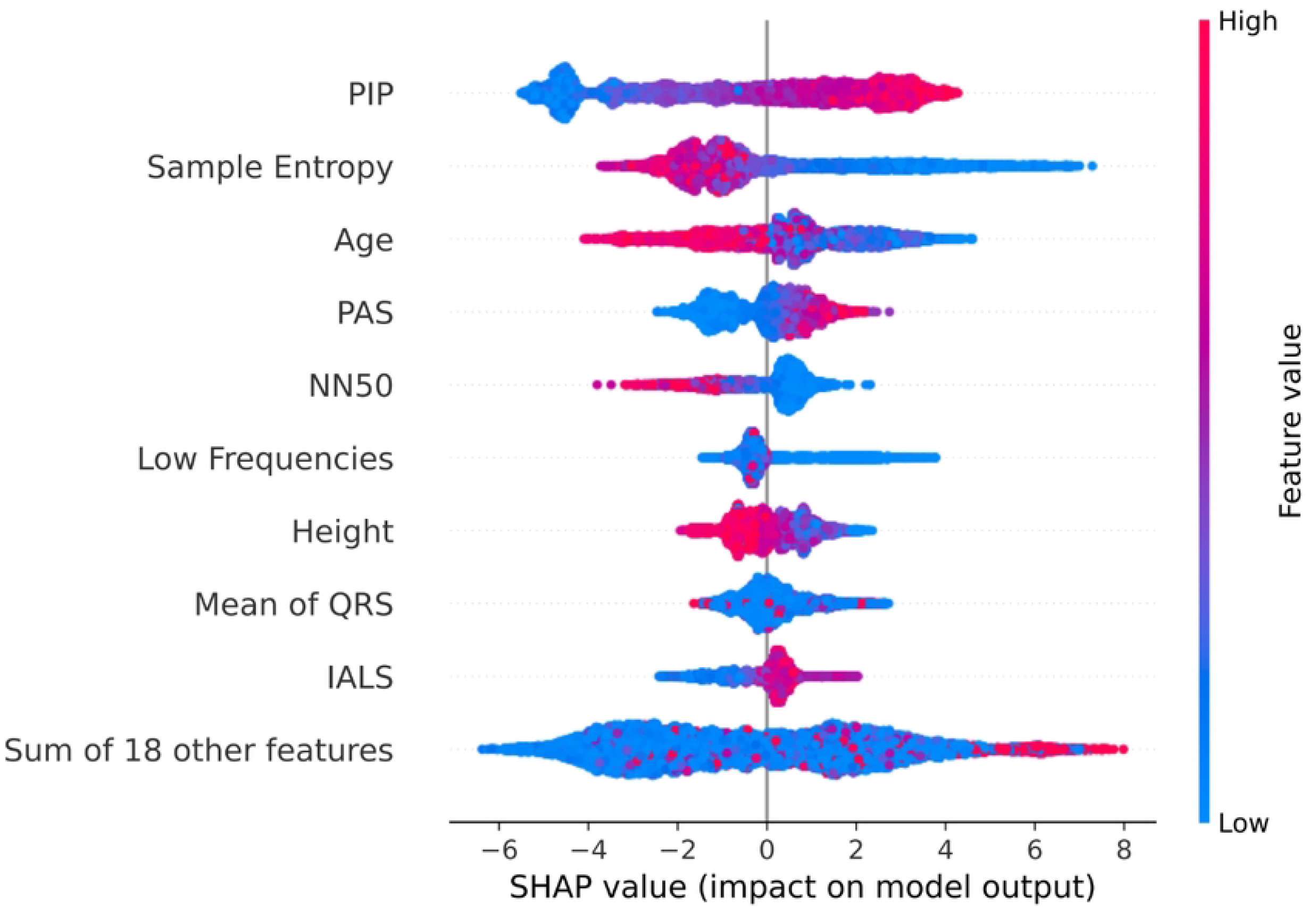
Features impact on XGBoost classification model predictions. In this plot a XGBoost model trained on the whole dataset was used for all calculations. Each point is colored (using a blue/low to red/high gradient) based on the value of one feature (y-axis) computed using one window of the dataset. Each point x-axis coordinate depicts the contribution of this point to the model output calculated using SHAP value. Positive SHAP values indicate an impact towards a positive (anesthetized) output. Likewise negative SHAP values indicate a contribution towards a negative (awake) output.

Patient characteristics such as age and to a lower extent height is the second category of features with the most importance for prediction. Fig 5 shows that age is the third most impactful feature on the model outcome.

Information dimension category is the third one per feature importance, mostly because the sample entropy is the second best discrimination feature as reported on Fig 5. Low sample entropy values impacts anesthetized state predictions on a very large range. On the other hand, high sample entropy values affect awake state predictions on the smaller scale. Notably, the other feature in this category, the permutation entropy, is not amongst the tenth features with the highest impact on the model predictions.

Morphological metrics, frequency domain HRV and in particular time domain HRV have a lesser influence on the model performances. In each of these feature categories a single feature has a significant impact on the model predictions as shown on Fig 5: mean of the QRS interval, low frequency HRV, and number of successive interbeat intervals longer than 50ms (NN50).

Consistent conclusions can be drawn from the the SVM and Random Forest feature importance analysis (see S1 Table). For the three classifiers considered, the percentage of inflection points (PIP), the sample entropy, and patient age importance account for one third of the total SHAP values. Furthermore, all HRF indices are in the top ninth most important features according to the average rank of these methods (see S1 Table).

## Discussion

Accurate assessment of depth of anesthesia is crucial for patient safety. This study demonstrates the potential of heart rate fragmentation (HRF) indices derived from electrocardiogram (ECG) signals to distinguish between awake and anesthetized states during general anaesthesia (GA). Data from 48 surgical patients, recorded both in the awake preoperative setting and during anesthesia, revealed significant differences in these metrics between the two states. While various features can be derived for the ECG, HRF and signal complexity indices associated with the patients’ physiological are decisive parameters for the performance of machine learning classifiers.

Our study demonstrated that a fraction of at least three minutes of ECG signal was optimal for the robustness of the classification models. Out of seven machine learning algorithms evaluated, the random forest, support vector machines and gradient boosting methods performed best (see Table 4) in distinguishing the awake and anesthetized states of our cohort. Still, the gradient boosting model demonstrated the best generalisation capability to discriminate anesthetized and non-anesthetized states using public datasets of asleep, awake, or anesthetized participants. Results of the feature importance analysis, calculated using Shapeley values, indicated that HRF metrics had the most significant impact on the classification performance. Additionally, patient age and sample entropy indices were identified as among the most contributing features. Conversely, frequency or time domain heart rate variability metrics had the lowest contribution on the classifier performances.

Classification of consciousness states is a challenging research topic [39]. Our understanding of the underlying biological and physical principles governing this complex physiological phenomenon remains limited [40]. Moreover, the precise mechanisms by which anesthetic agents induce reversible loss of consciousness remains to be elucidate [41]. In this study we used propofol to induced a loss of consciousness through potentiation of the inhibitory neurotransmitter *γ*-aminobutyric acid. This intravenous drug exerts well-documented effects on multiple organs, in particular the cardiovascular system [15, 42]. Previous research has established a correlation between states of consciousness under GA and heart rate variability (HRV) [17]. Early studies focus only on time and frequency domain HRV to discriminate the awake and anesthetized states [43, 44]. Our findings, however, demonstrate that these traditional HRV measures are less effective than HRF indices in discriminating these two states.

In their article Polk S.L. *et al*. [22] share the same goals as this study, develop an ECG-based system to discriminate between awake and anesthetized states. A total of twelve volunteers underwent anaesthesia using two anaesthetics setup: sevoflurane, and a combination of sevoflurane and ketamine. Best performances, evaluated by the F1 score, for the classification task were obtained using a logistic regression model: 0.83 for the sevoflurane (*resp*. 0.88 for the combined sevoflurane and ketamine) induced GA. The findings of this study demonstrate a performance outcome that is comparable to that of our own (0.84 for the gradient boosting model). Yet, there is less variability between participants and major experimental setup differences: the cohort was smaller (12 *vs* 48 in this study), volunteers were healthy and younger (mean age of 25±4.8 *vs* 51±16.2 in this study), no surgical procedure were performed, and the anaesthetic agents were different. Additionally, the methodological approach adopted by the authors exhibits notable divergences. In particular, the point-process method was employed for the probabilistic modelling of ECG signals, and only time and frequency domain HRV features were utilised, which proved to be less effective in the context of our study.

A recent study by Zhan J. *et al*. [23] investigated the potential of HRV metrics and sample entropy, combined with a deep neural network architecture, for classifying three consciousness levels during GA in twenty-three patients. These levels included induction (transition from wakefulness to anesthetized), maintenance of unconsciousness, and recovery of consciousness. Consciousness state labels were manually assigned by five expert clinicians. Methodological differences in the sampling process exist compared to our research. Zhan J. *et al*. used sliding windows of 256 RR intervals with a 5 RR overlap. Besides, our data showed that in most cases heart rate decreases below 60 bpm reflecting the observed bradycardia commonly encountered during general anesthesia. Consequently, Zhan J. *et al*. classification models were trained and evaluated on longer timescales (*e*.*g*. 256 sec at 60 bpm *vs* 180 sec in this study). This differences may contribute to the slightly accuracy improvement (0.90) compared to our results (0.84), considering that the classification task is not exactly the same. Notably, all classification models tested by Zhan J. *et al*. exhibited suboptimal performance in predicting both induction and recovery of consciousness, with precision scores ranging from 0.46 to 0.58. This limitation could be attributed to a potential class imbalance bias within the dataset, as the maintenance state typically constitutes the longest duration during surgical procedures. The balanced sampling procedure used in our study allows to mitigate such bias.

Both studies support the utility of ECG-derived features in distinguishing consciousness states, employing diverse cohort characteristics, experimental setups, and anesthetic agents. However, unlike our own work, these authors did not evaluate the generalizability of their approach.

Our study demonstrates that HRF indices play a significant role in enhancing the precision and reliability of the GA state classification task. These findings reinforce the relevance of these indices for this aim. Unlike HRV metrics, HRF indices are capable of distinguishing between vagally and non-vagally mediated heart rate fluctuations. Notably, in the context of short time scales and aging patients, fast heart rate fluctuations, which are not attributable to physiological vagal tone modulation, can impede the efficacy of traditional HRV analysis by inflating the values of short-term metrics such as root mean square of the successive differences and high-frequency power [24]. Several studies have shown a correlation between increased HRF metrics and advancing age [45], as well as a decline in signal complexity associated with aging [46]. Age is a well-established factor influencing cardiac function, demonstrably impacting various physiological metrics. ECG signals have been utilized to estimate both chronological age [47] and biological heart age [48], reflecting the cumulative effects of aging on cardiac health. Data presented herein confirm that HRF (*resp*. complexity) indices increase (*resp*. decrease) with the chronological age of awake patients (see S1 File, Fig 1 and Fig 2). Still, there is no correlation with age for anaesthetised patients. It can therefore be assumed that the ability of these metrics to discriminate between the two states is a limiting factor for elderly patients. However, as depicted in S1 File, HRF PIP metrics (Fig 1) is better as discriminating these two states than sample entropy (Fig 2), or time domain HRV RMSSD (Fig 3) and NN50 (Fig 4) indices. While these results suggest the potential of these metrics as anesthesia biomarkers, a study incorporating awake and anesthetized data from the same patients is crucial to mitigate inter-patient variability. Several limitations should be acknowledged when analyzing ECG signals, including their non-stationary nature and potential confounding factors. For instance, recordings during surgery are prone to more noise interference than those conducted in the pre-operative room. It is also subject to more interferences (*e*.*g*. electric scalpel, pulling out of electrodes). Accounting for the influence of these interferences, filtered in this study, could enable to develop robust models. Besides, Only propofol induced anesthesia has been considered. Further study is necessary to know if our results holds with other anesthetic agents.

The ECG signal is also used to monitor nociceptive pain. The underlying clinical hypothesis is that nociceptive pain alters the sympathetic/parasympathetic balance of the autonomic nervous system and indirectly the HRV. This raises the question of whether our models discriminate between the effect of analgesics and that of anesthetics. The HRV-based Analgesia/Nociceptive pain Index (ANI) from company Mdoloris (Lille, France) filters the normalised RR signal between 0.15 Hz and 0.50 Hz to retain only parasympathetic tone variations [49]. In contrast, our models are built without this filter. However, it would be interesting to compare ANI index to this study using analgesic/anaesthetic agents dose adjustments annotations. This question of causality between the different drugs used and cardiac activity during GA is a challenge for the research concerned.

To evaluate the generalizability of our models, we leveraged three public datasets: VitalDB, Fantasia, and MIT-BIH Polysomnographic. These datasets permit to assess performance across distinct consciousness states, with each focusing on a single state (anesthetized in VitalDB, awake in Fantasia, and asleep in MIT-BIH Polysomnographic). Our results demonstrated that both asleep (unseen during training of our models) and awake states were correctly classified as non-anesthetized. This finding highlights the specificity of our classifier in identifying the anesthetized state, particularly given the physiological similarities between asleep and awake states. Furthermore, the successful application of our model to the unfiltered ECG data within all datasets supports its robustness. However, it’s important to acknowledge limitations. The use of propofol as the sole anesthetic agent in VitalDB restricts the generalizability to other agents, particularly those targeting NMDA receptors like ketamine. Additionally, the single-state nature of each dataset prevents their direct utilization for training a GA classification task.

In this article, we demonstrated that it was possible to predict the state of consciousness (awake or anaesthetised) from the cardiac signal. The findings of this study provide a framework for the development of personalised indices for monitoring the depth of anaesthesia based on the ECG signal. To achieve this objective, it is essential to create cohorts tailored to the specific challenges of this research area. These cohorts should be sufficiently sizeable, multicentre, employ a diverse array of anaesthetic protocols and agents, and exhibit substantial inter-patient variability. Additionally, the development of methods for detecting transitions between different states of anaesthesia (*e*.*g*. from awake to induced) and the investigation of multimodal approaches represent promising avenues for future research, with the potential to enable the design of quantitative or semi-quantitative depth of anaesthesia indices.

## Data Availability

The data is owned by CHRU Tours Hospital and the authors do not have permission to share the data publicly as we are bound to the European General Data Protection Regulation (GDPR) as well as the subjects’ consent, stating that data may only be used for specific purposes and not be shared with 3rd parties. This is because the dataset comprises personal identifiable data, which not only holds for patients physiological parameters but also applies to electrocardiogram. That being clarified, there may be ways to make anonymized or minimized data available on requests. However, this must be governed by a data sharing and/or processing agreement, which limits the use of the data (e.g. only to consented purpose, with no attempts to re-identify subjects, etc.). The data is owned by: CHRU Tours Hospital, 2 Bd Tonnellé, 37000 Tours, France. In first instance, please contact

## Supporting information

**S1 Fig. Sample entropy upper bound plot**. These plots depict the sample entropy upper bound (detailed in Richman J.S. *et al*. [32]) as a function of the length of a time series. (A) sample entropy upper bound. (B) gradient of the sample entropy upper bound.

**S2 Fig. SVM performance metrics as a function of window size plot**. These plots depict four performance metrics (accuracy, F1 score, precision and recall scores) of a SVM classifier as a function of the segmentation window size (in seconds). The model was trained and evaluated on the whole dataset of this study.

**S3 Fig. SVM and random forest classifiers feature importance plots**.

(A)-(B) For each plot horizontal blue bars depict the mean of the absolute value of SHAP values per features groups and black lines the related standard deviation. For each classifier SHAP values are calculated using the following models trained on the whole dataset: (A) SVM classifier, (B) random forest classifier. (C)-(D) These beeswarm plots depict the feature impacts on the classification outcome of the following models: (C) SVM classifier, (D) random forest classifier. Each point is colored (using a blue/low to red/high gradient) based on the value of one feature (y-axis) computed using one window of the dataset. Each point x-axis coordinate depicts the contribution of this point to the model output calculated using SHAP value. Positive SHAP values indicate an impact towards a positive (anesthetized) output. Likewise negative SHAP values indicate a contribution towards a negative (awake) output.

**S1 Table. XGBoost, SVM and random forest classifiers SHAP values table**. This Excel file contains a single table with Shapeley values calculated for the three classifiers and all features. Three columns per classifier detail these calculations: the first one contains the SHAP value, the second one the percentage relative to the sum of SHAP values, and the third one the rank of the feature. Feature categories are indicated according to the background color.

**S1 File. HRF features performance evaluation**. This portable document file contains two tables and one figure. Tables report performance metrics of the seven models used in this study trained with different feature sets. Table 1 contains evaluation of models trained without heart rate fragmentation features. Table 2 contains evaluation of models trained using only heart rate fragmentation indices and patient characteristics. Figures 1-4 depict, as boxplots, the effects of aging in the awake and anesthetized states on the following metrics: Fig 1, percentage of inflection points (PIP); Fig 2: sample entropy (SampEn); Fig 3, root mean square of successive NNs differences; Fig 4, Number of pairs of successive NNs intervals ≥ 50 ms.

## Acknowledgments

This work has been supported as part of France 2030 programme, ANR-11-IDEX-003.

